# Two-step strategy for the identification of SARS-CoV-2 variant of concern 202012/01 and other variants with spike deletion H69-V70, France, August to December 2020

**DOI:** 10.1101/2020.11.10.20228528

**Authors:** Antonin Bal, Gregory Destras, Alexandre Gaymard, Karl Stefic, Julien Marlet, Sébastien Eymieux, Hadrien Regue, Quentin Semanas, Constance d’Aubarede, Geneviève Billaud, Frédéric Laurent, Claudia Gonzalez, Yahia Mekki, Martine Valette, Maude Bouscambert, Catherine Gaudy-Graffin, Bruno Lina, Florence Morfin, Laurence Josset, COVID-Diagnosis HCL Study Group

## Abstract

We report the implementation of a two-step strategy for the identification of SARS-CoV-2 variants carrying the spike deletion H69-V70 (ΔH69/ΔV70). This spike deletion resulted in a S-gene target failure (SGTF) of a three-target RT-PCR assay (TaqPath kit). Whole genome sequencing performed on 37 samples with SGTF revealed several receptor-binding domain mutations co-occurring with ΔH69/ΔV70. More importantly, this strategy enabled the first detection of the variant of concern 202012/01 in France on December 21^th^ 2020.

Since September a SARS-CoV-2 spike (S) deletion H69-V70 (ΔH69/ΔV70) has attracted increasing attention. This deletion was detected in the cluster-5 variant identified both in minks and humans in Denmark. This cluster-5 variant carries a receptor binding domain (RBD) mutation Y453F and was associated with reduced susceptibility to neutralizing antibodies to sera from recovered COVID-19 patients [1–3]. The ΔH69/ΔV70 has also co-occurred with two other RBD mutations of increasing interest [4]: N439K that is currently spreading in Europe and might also have reduced susceptibility to SARS-CoV-2 antibodies [5]; and N501Y that is part of the SARS-CoV-2 variant of concern (VOC) 202012/01 recently detected in England [6]. Although the impact of ΔH69/ΔV70 on SARS-CoV-2 pathogenesis is not clear, enhanced surveillance is urgently needed. Herein we report the implementation of a two-step strategy enabling a rapid detection of VOC 202012/01 or other variants carrying ΔH69/ΔV70.

## ΔH69/ΔV70 associated with S-gene target failure of a three-target RT-PCR assay

As part of routine SARS-CoV-2 genomic surveillance performed at national reference centre (NRC) for respiratory viruses (Lyon, France) [7], a 6-nucleotide deletion (21765-21770) within the S gene was identified in two nasopharyngeal samples collected on September 1^st^ and 7^th^, respectively. The SARS-CoV-2 infection diagnosis had been performed with the Applied Biosystems TaqPath RT-PCR COVID-19 kit (Thermo Fisher Scientific, Waltham, USA) that includes the ORF1ab, S, and N gene targets. For these two samples, a S-gene target failure (SGTF) was reported while ORF1ab and N targets were positive with Ct values < 25 (Figure 1A).

**Figure 1A.**
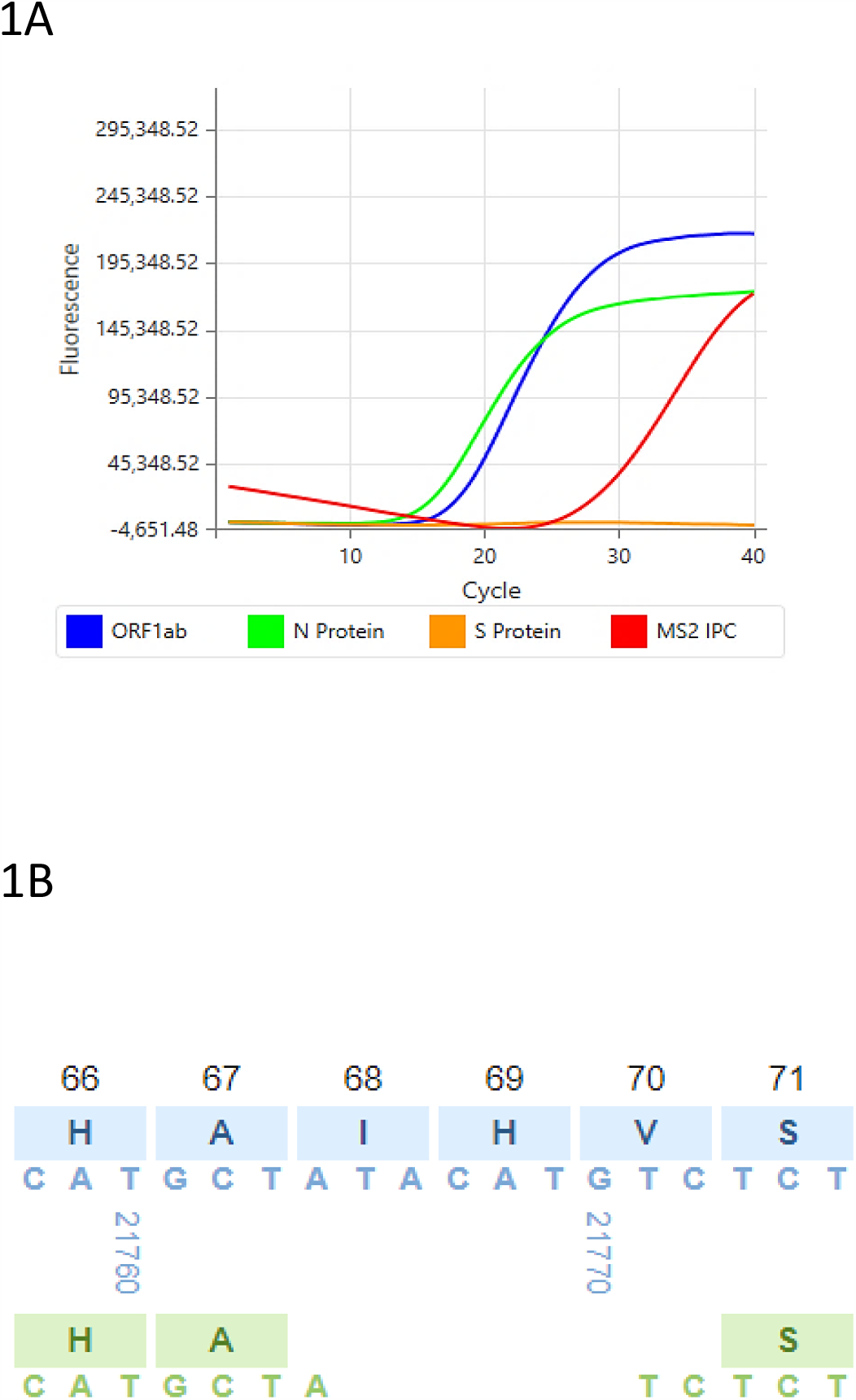
Amplification curves obtained with TaqPath COVID-19 RT-PCR kit for samples with the S deletion 21765-21770. The three targets included in the RT-PCR kit are represented by a different color. The amplification curve of the internal control is also represented (MS2, red curve). 1B. Pairwise sequence alignment from nucleotide position 21758 to 21775 of the spike gene using CoV-GLUE resource. Sequence with the deletion 21765-21770 is represented in green and the reference sequence in blue (Wuhan-Hu-1). The 21765-21770 deletion results in deletion of amino acid residues 69 and 70; ATC (21764-21771-21772) encoding for an isoleucine amino acid (I).

The mean coverage for the whole genome sequences generated was 6903x and 6898x, respectively and the S deletion 21765-21770 was present in 100% of the reads. Using CoV-GLUE online resource [8], we found that the S deletion 21765-21770 led to the removal of 2 amino acids (ΔH69/ΔV70) in the N-terminal domain of the S1 subunit of the S protein (Figure 1B). The whole genome sequencing (WGS) method used was the amplicon-based ARTIC v3 protocol (https://artic.network/ncov-2019) combined with Nextera DNA Flex library and sequencing on NextSeq 500 platform (Illumina, San Diego, USA). To confirm the presence of the deletion, one sample was also sequenced with an untargeted metagenomic protocol that yielded the same sequence. Of note, this metagenomic approach could not be applied for the second sample due to low viral load [9].

Although the coordinates of the primer/probe binding regions were not available for the TaqPath kit, the manufacturer confirmed that the S deletion H69-V70 was in the area targeted by the test.

## ΔH69/ΔV70 screening with RT-PCR followed by WGS

We then performed a retrospective analysis of RT-PCR results obtained using the TaqPath kit from August 3^rd^ to December 20^th^. We selected only positive samples with a Ct value < 25 for the N target, the most sensitive target of the test. By doing so, we found that 59/9,266 (0.6%) of positive tests had no amplification of the S gene. No significant increase of the SGTF was noticed over time; the proportion ranging from 0% (week # 32, 33, 34, 42, 48-51) to 2.91% (week # 35; Figure 2). Among the 59 samples with SGTF, 36 were available for WGS. These 36 samples were collected from August 5^th^ to November 11^th^ (18/36 were collected after October 9^th^). A total of 11 samples that presented an amplification of the S target were also sequenced. The sequencing results were fully concordant with the RT-PCR profiles (100% of the samples with SGTF had the S deletion ΔH69/ΔV70, while 100% of the S-gene positive samples did not contain ΔH69/ΔV70). For samples with SGTF, other S mutations were detected and are summarized in Table 1. The most frequent S mutations co-occurring with ΔH69/ΔV70 deletion were S477N and D614G that were found in 21/36 samples (58.3%). The co-occurrence of N439K and D614G mutations was found in 10/36 samples; the first sample containing this combination of mutation was collected on August 5^th^. Of note the complete combination of S mutations detected in cluster-5 variant was not found.

**Table 1.**
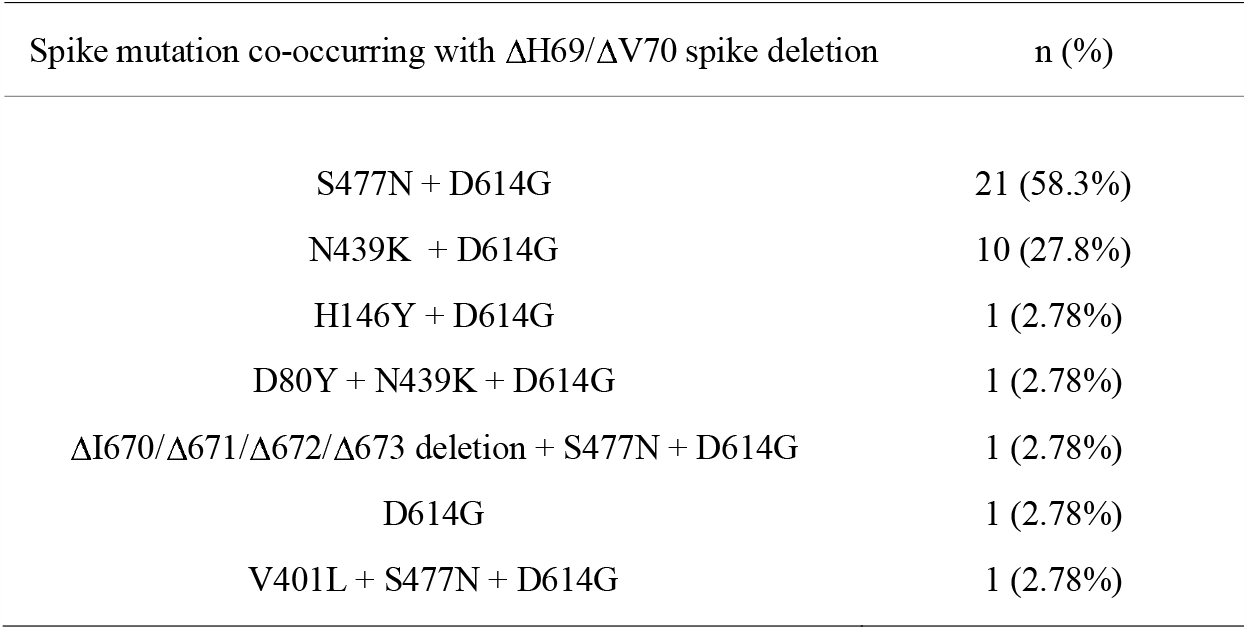
Spike mutations co-occurring with ΔH69/ΔV70 deletion in 36 samples with S negative profiles (negative for S target and positive for N & ORF1ab targets) obtained with the RT-PCR TaqPath kit.

| Spike mutation co-occurring with ΔH69/ΔV70 spike deletion | n (%) |
| --- | --- |
| S477N + D614G | 21 (58.3%) |
| N439K + D614G | 10 (27.8%) |
| H146Y + D614G | 1 (2.78%) |
| D80Y + N439K + D614G | 1 (2.78%) |
| ΔI670/Δ671/Δ672/Δ673 deletion + S477N + D614G | 1 (2.78%) |
| D614G | 1 (2.78%) |
| V401L + S477N + D614G | 1 (2.78%) |

**Figure 2.**
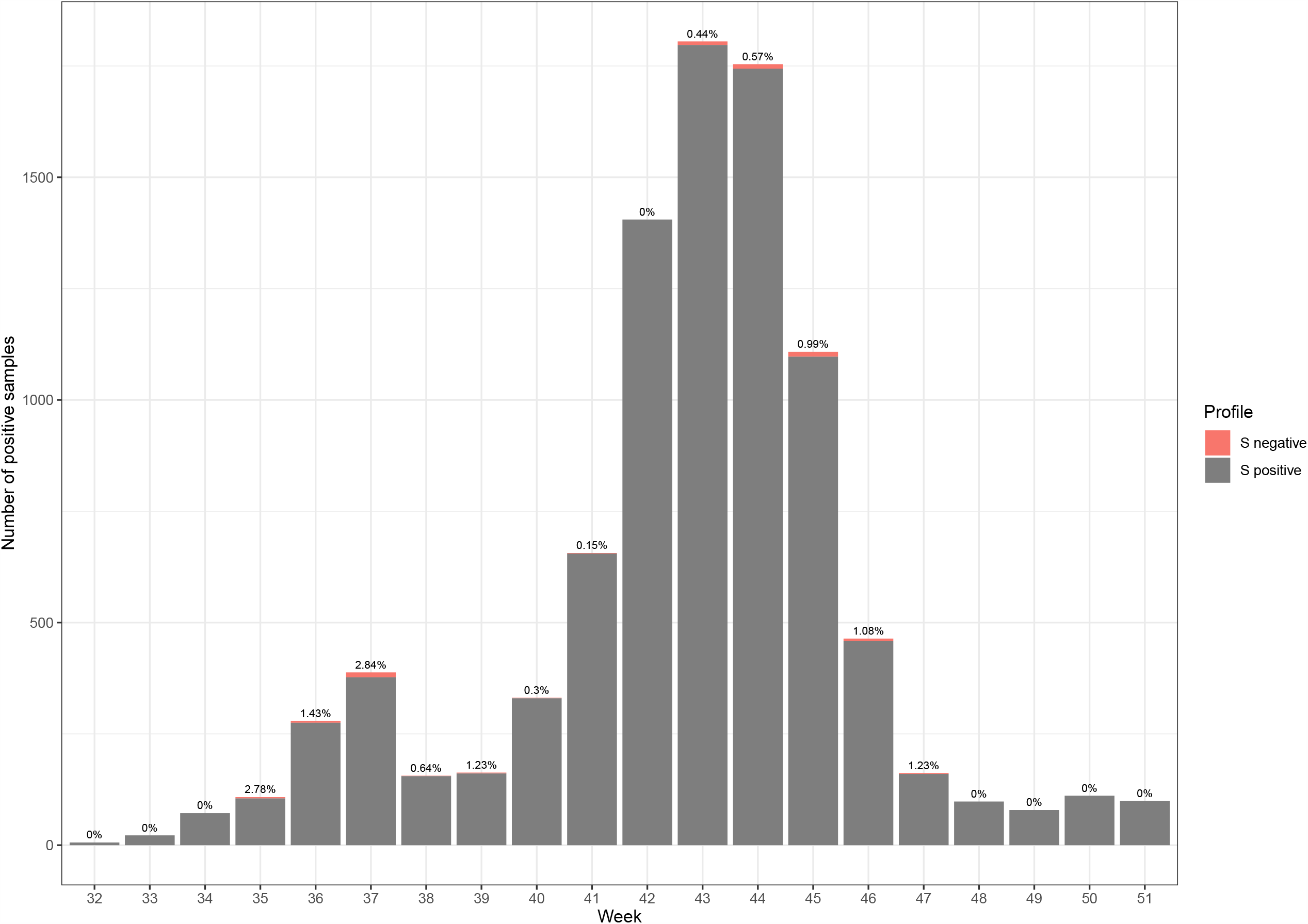
Prevalence of the S negative profile (negative for S target and positive for N & ORF1ab targets) with TaqPath COVID-19 RT-PCR kit from August 3^rd^ (week 32) to December 20^th^ (week 51).

The 2-step strategy presented herein and based on a screening with TaqPath Kit followed by WGS for samples with SGTF has been implemented in France since December 20^th^. On December 21^th^, the virology laboratory of university hospital of Tours reported a SGTF on a nasopharyngeal sample from a patient with a recent travel history from England (London). The sample was addressed to NRC for WGS and the detection of VOC 202012/01 (lineage B.1.1.7) was confirmed on December 25^th^ that corresponded to the first detection of this variant in France (GISAID accession number EPI_ISL_735391).

## Discussion and conclusion

According to CoV-GLUE resource [8] (last update from GISAID: December 14^th^), the S deletion 21765-21770 has been identified in 4,632 sequences worldwide (>99% in Europe) Interestingly, only 16 sequences containing this deletion were sampled between March 15^th^ and July 23^th^ corresponding to the first wave of COVID-19 pandemic in Europe. Herein, using data obtained with TaqPath RT-PCR kit, we found an overall prevalence of 0.6%, suggesting a limited circulation of variants presenting the ΔH69/ΔV70 deletion during the second wave of the pandemic in Lyon, France.

It should be underlined that N439K, Y453F, or N501Y RBD mutations that can co-occurred with ΔH69/ΔV70 deletion might be associated with an increased affinity to ACE2 or reduced sensitivity to SARS-CoV-2 antibodies [3, 5, 10–12]. It has been hypothesized that the ΔH69/ΔV70 deletion might compensate some RBD mutations and might be involved in the transmissibility of variant containing these mutations [4, 6]. In addition, it has been recently shown that the combined ΔH69/V70 and D796H mutant was less sensitive to neutralizing antibodies [13]. As the N-terminal domain may interact with lung receptors [14] and might be a target of neutralizing antibodies [15, 16], further studies are needed to understand the consequences of ΔH69/ΔV70 deletion on SARS-CoV-2 transmissibility and host-immune response.

Importantly, the TaqPath kit used for this study did not lead to a false negative conclusion as the two other targets remain positive. The data presented herein emphasize that the TaqPath RT-PCR assay is a useful and cost-effective tool enabling a rapid, large-scale screening of SARS-CoV-2 variants with ΔH69/ΔV70. Samples with SGTF should be further addressed to national referral laboratories for SARS-CoV-2 WGS. This 2-step strategy can contribute to the early detection of SARS-CoV-2 VOC 202012/01 which has been found to be more transmissible than non-VOC lineage [17]. This strategy is currently being reinforced in France as national diagnostic platforms have mainly implemented the TaqPath RT-PCR kit.

## Data Availability

SARS-CoV-2 genomes sequenced in this study were deposited in the GISAID database (EPI_ISL_582110, EPI_ISL_582111, EPI_ISL_582112, EPI_ISL_582113, EPI_ISL_582114, EPI_ISL_582115, EPI_ISL_582116, EPI_ISL_582117, EPI_ISL_582118, EPI_ISL_582119, EPI_ISL_582120, EPI_ISL_582508, EPI_ISL_623098, EPI_ISL_623099, EPI_ISL_623100, EPI_ISL_623101, EPI_ISL_623102)

## Data availability

SARS-CoV-2 whole genomes sequenced in this study were deposited in the GISAID database (EPI_ISL_582110, EPI_ISL_582111, EPI_ISL_582112, EPI_ISL_582113, EPI_ISL_582114, EPI_ISL_582115, EPI_ISL_582116, EPI_ISL_582117, EPI_ISL_582118, EPI_ISL_582119, EPI_ISL_582120, EPI_ISL_582508, EPI_ISL_623098, EPI_ISL_623099, EPI_ISL_623100, EPI_ISL_623101, EPI_ISL_623102, EPI_ISL_735391)

## Ethics statement

Samples used in this study were collected as part of an approved ongoing surveillance conducted by the national reference centre for respiratory viruses in Lyon, France (WHO reference laboratory providing confirmatory testing for COVID-19). The investigations were carried out in accordance with the General Data Protection Regulation (Regulation (EU) 2016/679 and Directive 95/46/EC) and the French data protection law (Law 78–17 on 06/01/1978 and Décret 2019–536 on 29/05/2019). Samples were collected for regular clinical management during hospital stay, with no additional samples for the purpose of this study. Patients were informed of the research and their non-objection approval was confirmed. This study was presented by the ethics committee of the Hospices Civils de Lyon (HCL), Lyon, France and registered on the HCL database of RIPHN studies (AGORA N°41).

